# Predicting intention to donate blood among secondary school students in Eastern Uganda: An application of the theory of planned behavior

**DOI:** 10.1101/2024.06.20.24309241

**Authors:** Martha Akulume, Angela N Kisakye, Florence R Nankya, Suzanne N Kiwanuka

## Abstract

**Introduction:** The availability of donated blood in Uganda rests on the good will of voluntary blood donors. To design interventions aimed at improving the availability of donated blood, there is need to understand the predictors of blood donation. Previous studies have shown that intention to donate blood predicts the practice of blood donation.

**Aim:** This study aimed at applying the theory of planned behaviour to predict intention to donate blood among secondary school students, the major blood donor group in Uganda.

**Methods:** We conducted a cross-sectional study among 336 students from four purposively selected secondary schools in Eastern Uganda. Census sampling was used to recruit the students for this study. A self-administered questionnaire was used to collect data on socio-demographic characteristics as well as the theory of planned behavior constructs. Data were analyzed in STATA version 14 using descriptive statistics and regressions.

**Results:** About 75% (n=252) of the students had intention to donate blood sometime in their life. Students’ perceptions of their ability to donate blood (perceived behavioural control) was the key predictor of intention to donate blood (AOR = 6.35, CI =3.32, 12.15). The influence of subjective norms (AOR = 1.97, CI = 0.92, 4.20) and attitudes towards blood donation (AOR = 1.51, CI = 0.80, 2.87) did not significantly influence blood donation adjusting for other theory of planned behaviour constructs. The theory of planned behaviour constructs explained 15.5% of the students’ intention to donate blood. Regarding the external factors, only location of the school (AOR = 0.50, CI = 0.27, 0.93) and knowing someone who has ever donated (AOR = 0.26, CI = 0.12, 0.56) predicted the intention to donate blood.

**Conclusion:** Most students intended to donate blood sometime in their life. Efforts should be made to make blood donation sites accessible to students.

## Introduction

Globally, millions of lives are saved through blood transfusion each year. Despite this, the availability of blood for transfusion remains a challenge in developing countries, especially in rural areas [1, 2]. Although the World Health Organisation (WHO) recommends that at least 1% of the country’s population should donate blood in order to meet the essential blood donation needs [3], only 0.5% of the population in sub-Saharan countries voluntarily donate blood [4, 5]. As such, many of the sub-Saharan countries are able to collect only half of blood required to meet their populations needs [6].

Uganda, one of the sub-Saharan countries requires at least 340,000 units of safe blood each year. Despite this, the country collects only 200,000 units annually [7]. To address the challenges of blood shortages in Uganda, a number of interventions have been implemented in the country the past decade, including; the establishment of seven regional blood banks that collect, test, and store blood as well as partnerships between ministry of health and various organisations (for example; the Uganda red cross society, the Uganda blood transfusion services and voluntary blood donor clubs) to host blood donation drives [8, 9]. Despite these developments, there are not enough blood donations to satisfy the medical demand for blood in the country [9]. Considering that lack of access to blood transfusions leads to an increase in the number of preventable deaths [9], there is an urgent need for interventions to address blood shortages in the country.

In most developing countries such as Uganda where there are no regular blood donors, blood availability is at the mercy of voluntary individuals, who are the main source of blood [10]. Moreover, it is estimated that 90% of the blood donors in Uganda are secondary school students [11]. Understanding the predictors of their blood donation behaviour is the first and key step in designing interventions aimed at improving blood donations. Previous studies have shown that intention (readiness) to donate blood predicts the practice of blood donation [12, 13]. According to Ajzen’s theory of planned behaviour (TPB), intention to donate blood is further determined by a person’s attitude towards blood donation (feeling of favourableness or unfavourableness towards blood donation), the influence of subjective norms (perceived social pressure to donate blood) and perceived behavioural control (a person’s perception of his or her ability to donate blood) [14, 15].

The TPB is as an expansion of the Theory of Reasoned Action (TRA) [16]. The TRA postulated that intention to perform a certain behaviour was determined by the attitude towards a behaviour and the influence of significant others [16]. However, in circumstances where a person doesn’t have full control over their behaviour, behaviour cannot be determined exclusively by their intention to perform the behaviour. Therefore, to address the limitations of the TRA, the TPB was introduced by adding a new concept, i.e. perceived behavioural control [16]. Perceived behavioural control comprises of the internal and external factors that influence behaviour either directly or indirectly through intentions [17].

The TPB has been increasing applied to determine blood donation behaviours [10, 18, 19]. Nonetheless, Parash and others note that most of these studies are conducted in the Indian and Western context [20]. This study therefore applies the TPB to predict blood donation behaviours among secondary school students in Uganda.

## Methods

### Study setting and population

This was a cross sectional study conducted among students in four secondary schools in Eastern Uganda (i.e. two schools in Jinja and two schools in Mayuge districts) as part of the *“blood donation awareness project: a quasi-experimental”* study end-line survey [21]. Data were collected in April, 2022. The study included senior four and senior six students from the four schools because they had been involved in the earlier stages of the “*blood donation awareness project: a quasi-experimental”* study (the base-line survey and implementation of the intervention). Of the 559 students, who consented to participate in the end-line interviews, only 336 completed the questions related to the TPB constructs leading to a response rate of 60.1%.

### Sampling and procedure

We purposively selected two study districts i.e. Jinja city to represent an urban district and Mayuge district to represent a rural district. At district level, we worked with the district education officers to purposively select two secondary schools within each district to represent a highly populated and low populated school. While at the schools, census sampling was used for the survey and all the senior four and senior six students were invited to participate in the study. At each school, we called for an assembly where we informed the students about the study, its voluntary nature, purpose and procedures. All the interested students were given self-administered questionnaires.

### Questionnaire

Questions on the TPB constructs were included in the end-line survey questionnaire for the “*blood donation awareness project: a quasi-experimental”* study. The questionnaire was developed in English and consisted of 59 items to assess the TPB constructs.

#### Intention

Intention to donate blood was assessed using four items for instance *“I intend to donate blood in the next blood donation campaign/ event at my school”*. Responses were measured on a five point Likert scale, ranging from 1 (strongly agree) to 5 (strongly disagree). The Cronbach’s alpha for this scale was 0.760.

#### Perceived behavioral control

Six items were included to measure perceived behavioral control, for instance *“My school supports me to donate blood”.* The items were measured on a five point Likert scale, ranging from 1 (strongly disagree) to 5 (strongly agree). The Cronbach’s alpha for this scale was 0.717.

#### Subjective norms

To assess the influence of subjective norms on intention to donate blood, we included ten items, for instance *“My classmates think that blood donation is important”*. Responses were measured on a five-point Likert scale, ranging from 1 (strongly agree) to 5 (strongly disagree). The Cronbach’s alpha for this scale was 0.771.

#### Attitudes

Seven items were used to assess attitudes towards blood donation, for example *“Blood donation is a waste of time”*. Responses were measured on a five point Likert scale, ranging from 1 (strongly agree) to 5 (strongly disagree). The Cronbach’s alpha for this scale was 0.765.

#### External factors

In addition to the TPB constructs, the self-administered questionnaire collected data on; the socio-demographic characteristics (such as; age, sex, academic level etc.), knowledge and practices regarding blood donation.

#### Pretesting

The questionnaire was pre-tested among 30 secondary students from a school located in Kampala prior to data collection. The data from the pre-test was not included in the final analysis but was used to inform adjustments in the tool.

### Data Management and analysis

Data were entered using Microsoft Access and later exported to STATA 14 for analysis. First, the distributions of each variable were inspected to check for errors. Reliability tests were then performed for each TPB construct using the Cronbach’s alpha. The Likert scales for each TPB construct (intention, perceived behavioural control, attitude, and subjective norms) were computed. To obtain each participants’ overall score for the individual TPB constructs, we obtained the mean score all items assessing each construct. The new variable was categorized in five levels i.e.1 (strongly agree) to 5 (strongly disagree). The outcome variable intention was computed into a binary variable, categorized as “yes” (Strongly agree and agree) and “no” (strongly disagree, disagree and neither agree nor disagree). Descriptive statistics were used to investigate participants’ characteristics. Beyond descriptive statistics, associations were analysed using correlations and regressions.

### Ethical approval

We obtained ethical approval from Makerere University, School of Public Health Research and Ethics Committee (protocol no.852). Permission to conduct research was obtained from the Uganda National Council of Science and Technology (protocol no. HS1243ES) while administrative clearance was obtained from the district education officers and school head teachers. The students were required to provide their informed consent before participating in the study. For those aged <18 years, we obtained parental consent and assent from the students. The school administration was requested to invite the parents of the students aged <18years. We explained the purpose of the study to the parents and what it entailed. The parents were then asked to provide consent for their students to participate in the study.

## Results

### Participants’ characteristics

Slightly more than half of the respondents were male (n=188, 56.3%) while the mean age of the respondents was 19.0 years (SD = 2.2). Nearly half of the students (n=169, 50.3%) were from schools located in rural areas while majority of them had access to internet (n=237, 71.0%). Regarding their knowledge about blood donation, slightly less than half of the students (n=164, 48.8%) were knowledgeable about blood donation (Table 1).

**Table 1.**
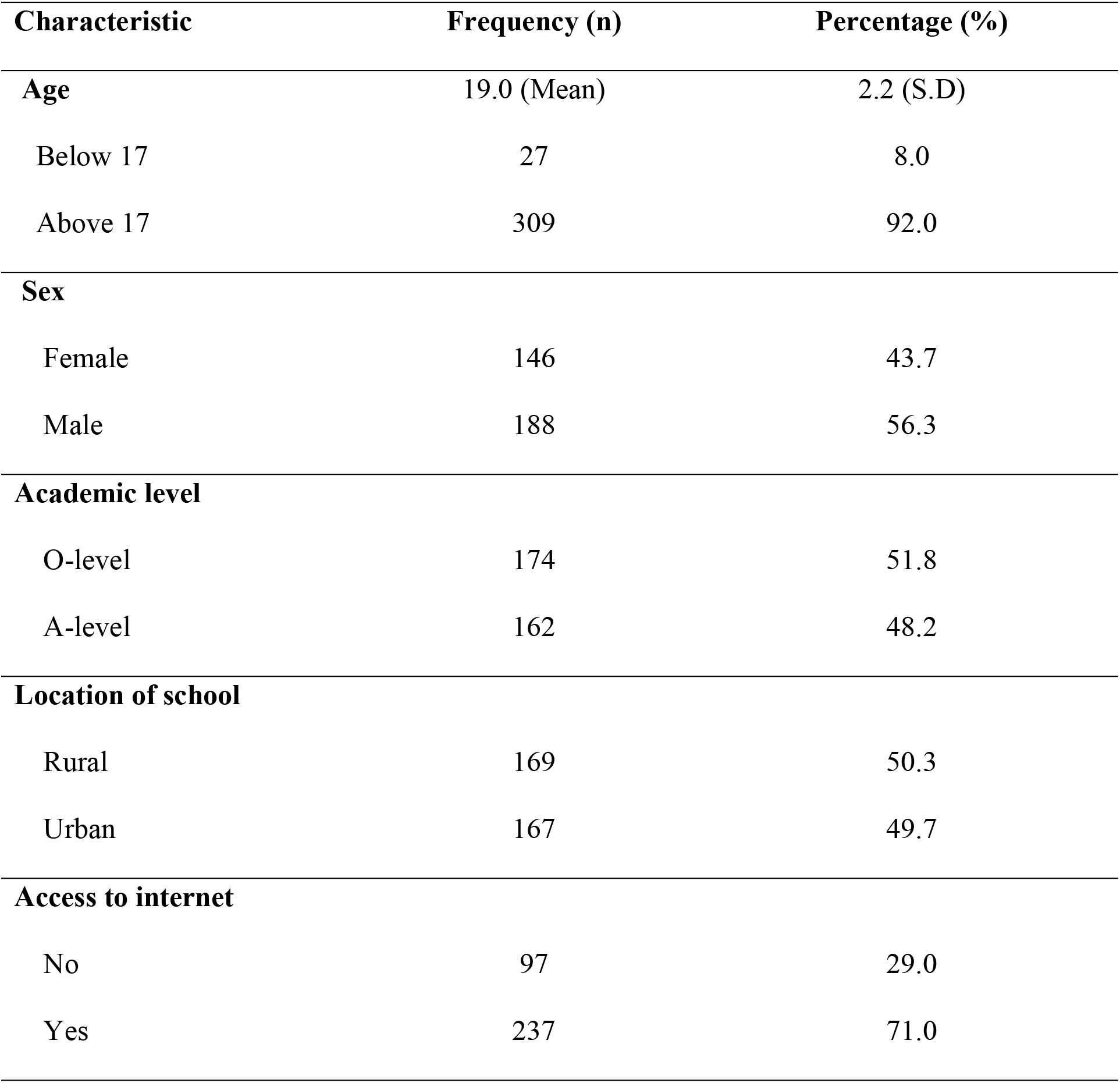
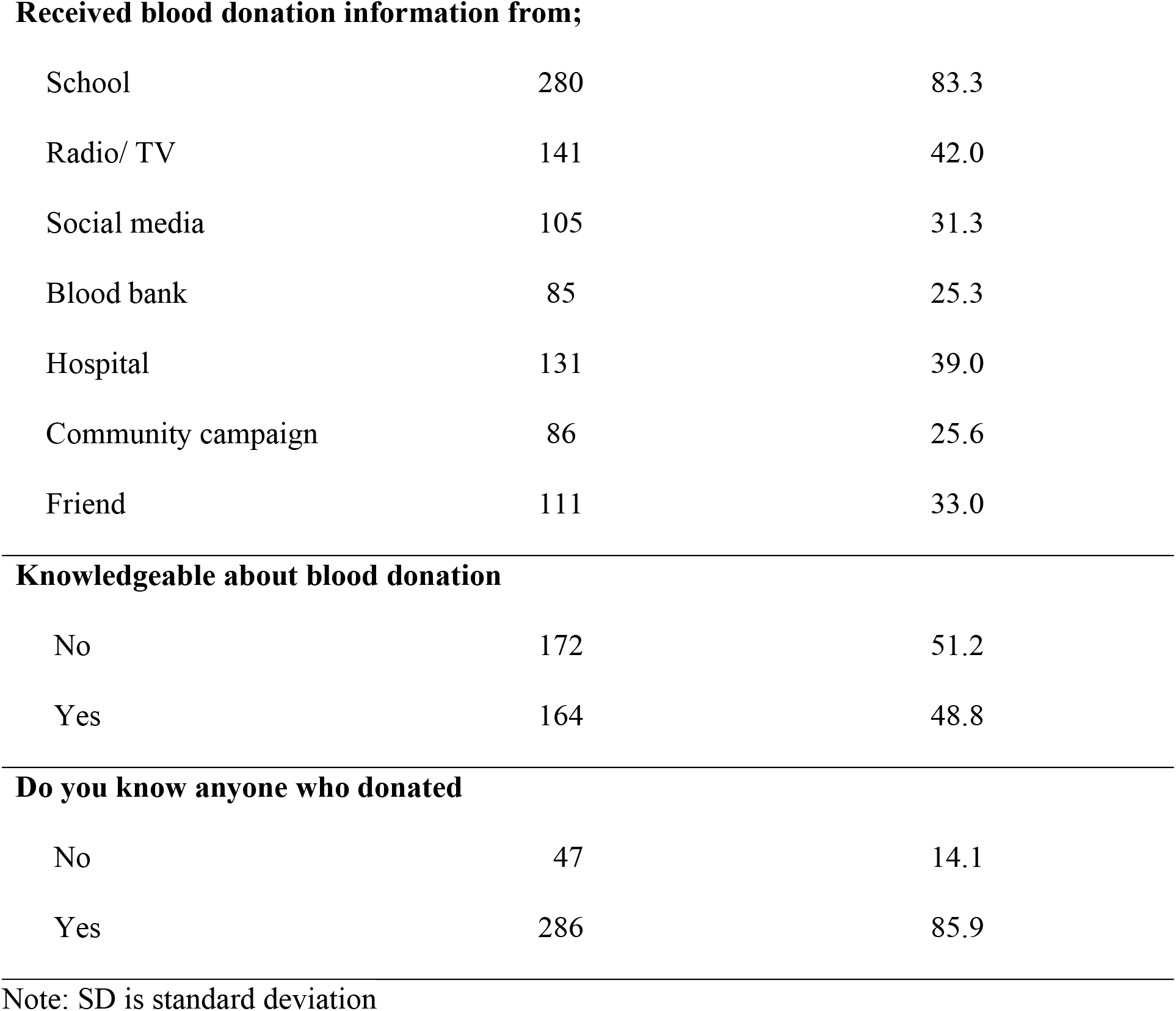
Characteristics of participants.

| Characteristic | Frequency (n) | Percentage (%) |
| --- | --- | --- |
| **Age** | 19.0 (Mean) | 2.2 (S.D) |
| Below 17 | 27 | 8.0 |
| Above 17 | 309 | 92.0 |
| **Sex** |  |  |
| Female | 146 | 43.7 |
| Male | 188 | 56.3 |
| **Academic level** |  |  |
| O-level | 174 | 51.8 |
| A-level | 162 | 48.2 |
| **Location of school** |  |  |
| Rural | 169 | 50.3 |
| Urban | 167 | 49.7 |
| **Access to internet** |  |  |
| No | 97 | 29.0 |
| Yes | 237 | 71.0 |
| **Received blood donation information from;** |  |  |
| School | 280 | 83.3 |
| Radio/ TV | 141 | 42.0 |
| Social media | 105 | 31.3 |
| Blood bank | 85 | 25.3 |
| Hospital | 131 | 39.0 |
| Community campaign | 86 | 25.6 |
| Friend | 111 | 33.0 |
| **Knowledgeable about blood donation** |  |  |
| No | 172 | 51.2 |
| Yes | 164 | 48.8 |
| **Do you know anyone who donated** |  |  |
| No | 47 | 14.1 |
| Yes | 286 | 85.9 |

Note: SD is standard deviation

### Intention to donate blood

About 75% of the students had intention to donate blood sometime in their life time [strongly agree - 33.3% (n= 112) and agree - 41.7% (n= 140)] while 25% did not intend to donate blood at any time in their life (See Fig 1).

**Fig 1.**
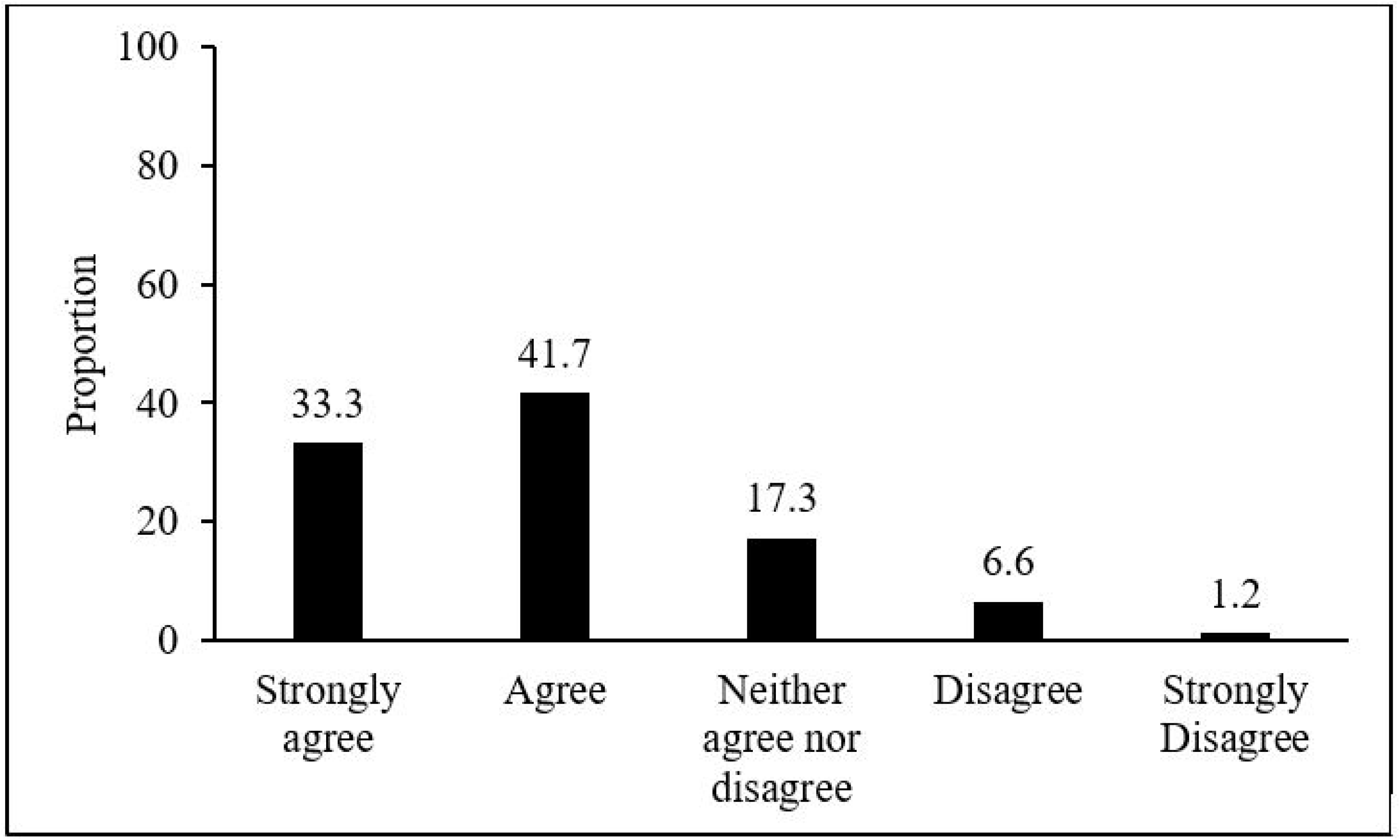
Students’ intention to donate blood

### Association between TPB constructs

As shown in Table 2, significant correlations were found between students’ intention to donate blood and the other TPB constructs, with perceived behavioural control emerging as the strongest correlate of intention to donate (r = 0.496, P value < 0.05). The matrix also revealed a moderate correlation (r = 0.450, P value < 0.05) between influence of subjective norms and intention to donate blood. Attitudes had a low correlation with intention to donate (r = 0.328, P value < 0.05)

**Table 2.** Correlations Between The TPB Constructs.

| Variable | Intention | PBC | Attitude | Subjective norms |
| --- | --- | --- | --- | --- |
| Intention | 1 |  |  |  |
| PBC | 0.496* | 1 |  |  |
| Attitude | 0.328* | 0.342* | 1 |  |
| Subjective norms | 0.450* | 0.504* | 0.483* | 1 |

Note: PBC is perceived behavioural control

### Significance of the TPB in predicting intention to donate blood

As shown in Table 3, the TPB constructs (attitudes, subjective norms and Perceived behavioural control) only explained 15.5% of the student’s intention to donate blood. Perceived behavioural control (AOR = 6.35, CI = 3.32, 12.15) was the main predictor of students’ intention to donate blood (See Table 3(.

**Table 3.** Regression analyses of TPB constructs.

| Variables | COR [95% CI] | P-Value | AOR [95% CI] | P-Value |
| --- | --- | --- | --- | --- |
| **PBC** | **8.64 [4.74, 15.74]** | **0.000** | **6.35 [3.32, 12.15]** | **0.000** |
| Attitude | 2.49 [1.45, 4.26] | 0.001 | 1.51[0.80, 2.87] | 0.206 |
| Subjective norms | 4.75 [2.56, 8.82] | 0.000 | 1.97 [0.92, 4.20] | 0.080 |

Note: R-Square for the multivariable model = 0.155
PBC is perceived behavioural control

### Relationship between perceived behavioural control and intention to donate blood

Further analysis showed that the student’s perception about their ability to donate blood (AOR = 2.42, CI = 1.77, 3.29), support from the school (AOR = 1.48, CI = 1.12, 1.97) and support from other stakeholder involved in blood donation (AOR = 1.43, CI = 1.07, 1.90) were the key domains of perceived behavioural control that influenced student’s intention to donate blood. (Table 4).

**Table 4.**
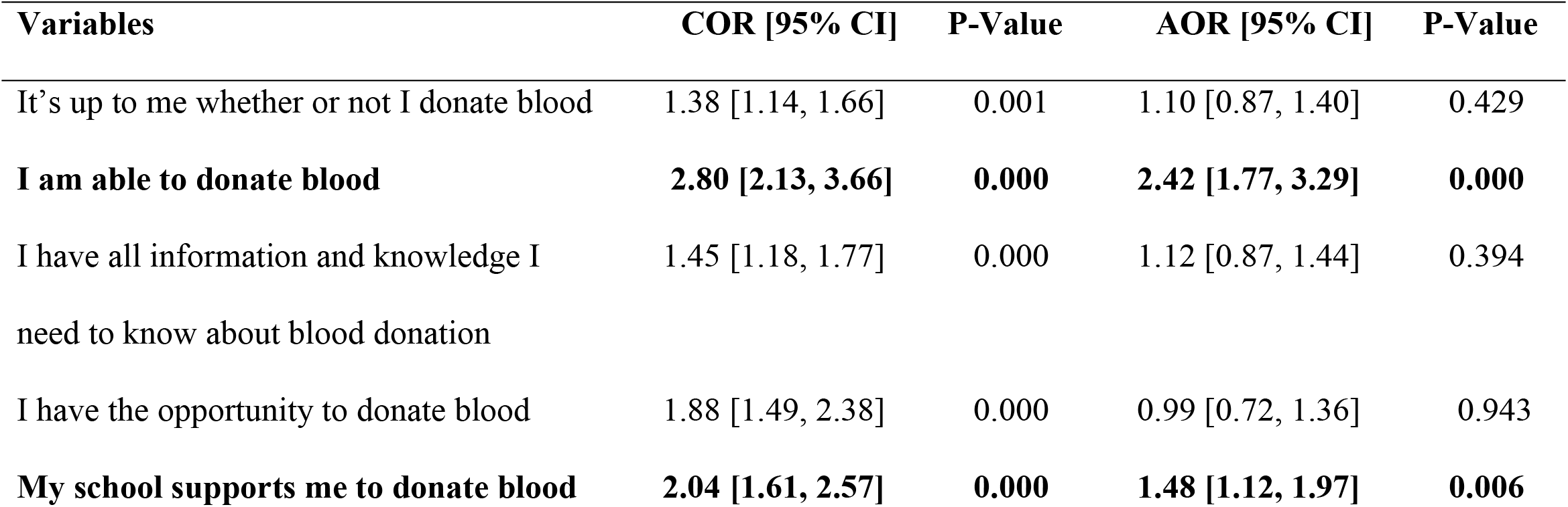
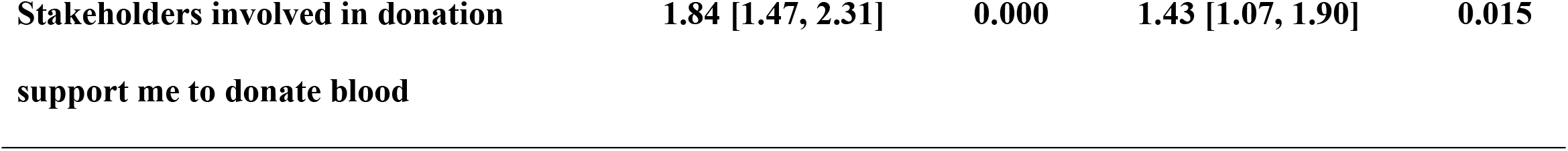
Regression analysis of perceived behavioural control and intention to donate blood.

**Table 5.**
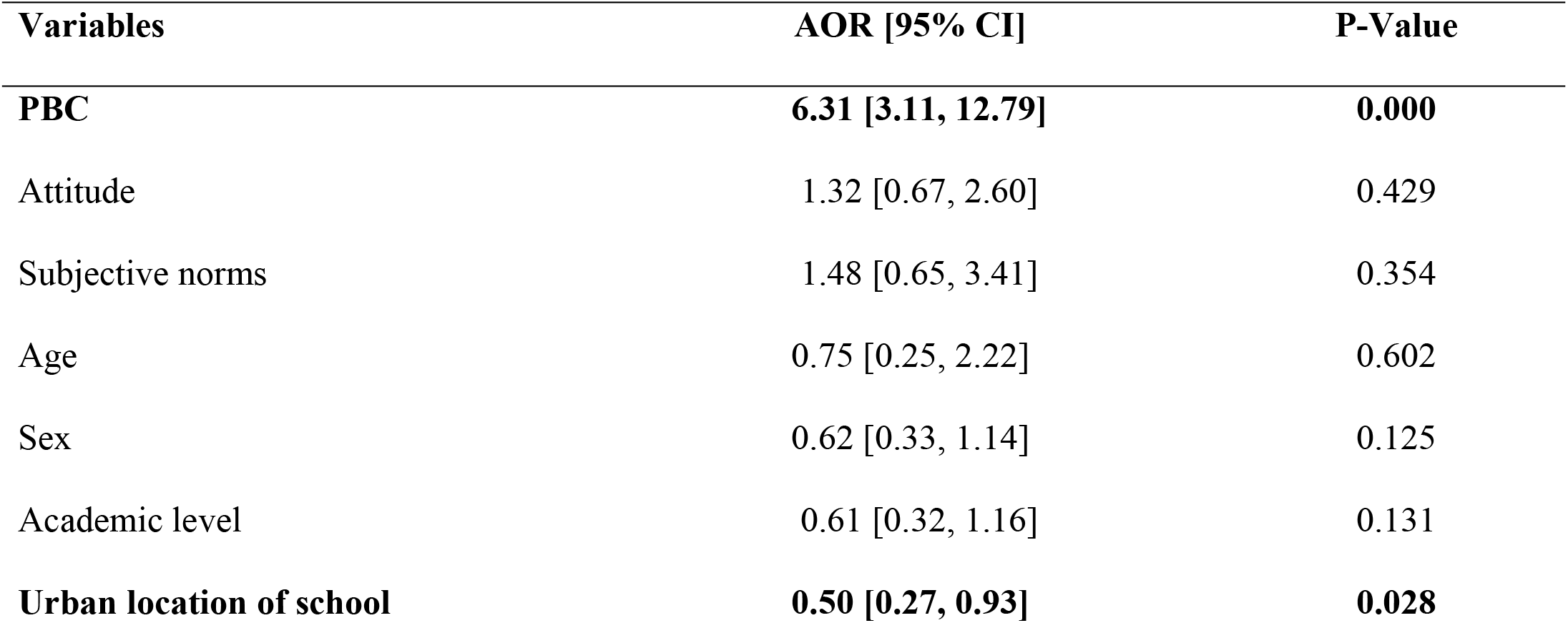
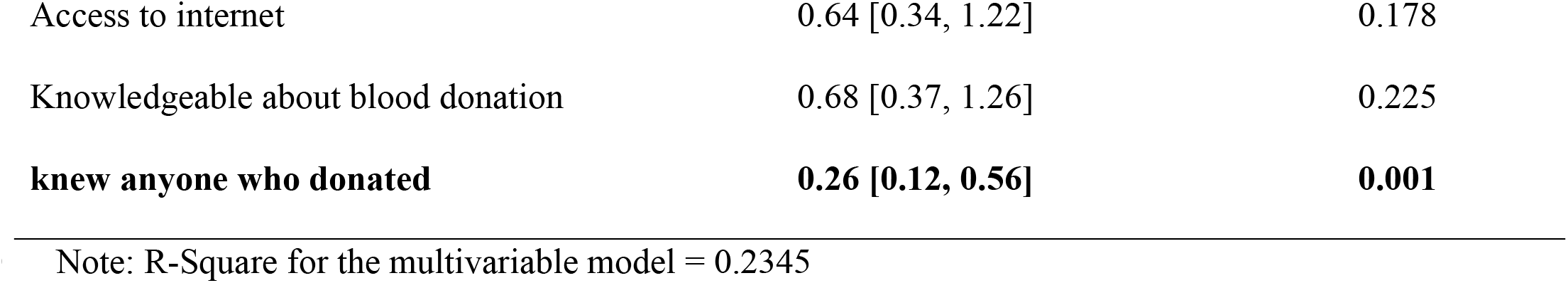
Multivariable regression analysis of TPB constructs adjusting for socio-demographic variables to predict intention to donate blood.

### Significance of external variables and the TPB Constructs in predicting students’ intention to donate blood

When external variables were added to the model, 23.5% variance in the students’ intention to donate blood was explained by the new model. The multivariable model also shows that perceived behavioural control (AOR = 6.31, CI = 3.11, 12.79) was the only TPB construct significantly associated with intention to donate. Additionally, students from an urban school (AOR = 0.50, CI = 0.27, 0.93) and those who knew someone who had ever donated blood (AOR = 0.26, CI = 0.12, 0.56) were likely to have low intentions to donate.

## Discussion

We aimed at applying the TPB to determine students’ intention to donate blood. Our findings showed that majority of the students intended to donate blood at some point in their life. This finding is similar to that of a previous study conducted among residential students and their teachers in Bangladesh which found that majority of the respondents were willing to donate blood to an unknown person [22]. This could be attributed to the fact that donating blood can potentially save lives [23] and therefore, most people are willing to donate so as to save lives. Considering that students already have a high intention to donate blood, it is therefore important that they have the appropriate information regarding blood donation and are also provided with avenues to donate blood. Another study conducted among private higher education students in Ethiopia showed that the mean intention to donate blood was low [24]. The difference between the two studies could be attributed to the fact that the present study was conducted after a blood donation awareness campaign [21] which could have influenced the students’ willingness to donate.

This study further showed that the TPB was a weak predictor of the student’s intention to donate blood. This finding differs from those of a study conducted among the students in Ethiopia where the TPB explained more than half of the students’ intention to donate blood [24, 25]. This could be attributed to the difference in the study areas. Among the TPB constructs, only perceived behavioural control significantly predicted the students’ intention to donate blood after adjusting for other TPB constructs. The perceived behavioural control domains that were significantly associated with blood donation were perceived support from school and other stakeholders involved in blood donation. Other studies have also found PBC to be associated with the intention to donate blood [10, 24]. This finding implies that students who intend to donate blood in their life time are those who are confident enough to donate [10]. Therefore, the Uganda blood transfusion services should work with the school administrations to make the process of blood donation and the environment where donation is carried out more conducive so as to boost student’s confidence to donate blood.

This study showed that the influence of subjective norms and the attitude towards blood donation did not significantly predict students’ intention to donate blood. Incongruent with this finding, Kassie and others [10], found subjective norms and attitude domains to be significant predictors of blood donation intentions. The difference between the two studies could be attributed to the difference in the study populations. Whereas the present study was conducted among secondary school students, the former study was conducted among adults in the community. Nonetheless, Faqah et al. [25], also found that subjective norms did not influence students’ intention to donate blood. This may imply that for altruistic behaviours like blood donation, significant others do not influence the intention to perform a behaviour but rather an individuals’ perception of their confidence to perform a behaviour.

The variance explained by the model increased when external factors were added to the model, implying that external factors (such as; social demographic characteristics and knowledge about blood donation) are key predictors of blood donation behaviours. The study further showed that the students who were in urban schools had a lower intention to donate blood compared to those in rural areas. This finding differs from that of a study conducted in Nigeria to assess rural-urban variation in willingness to donate blood which showed that urban dwellers were more willing to donate blood than rural dwellers [26]. This finding could be attributed to the difference in the socio-demographic characteristics between rural and urban students or could be due to lack of awareness of the blood donation sites. Therefore, the ministry of health should continue partnering with other institution to create awareness of the different blood donation sites.

Surprisingly, the students who knew someone who had donated had a lower intention to donate blood compared to those who did not know anyone who had donated. Again, this finding may imply that for altruistic behaviours like blood donation, significant others do not influence the intention to perform a behaviour but rather an individuals’ perception of their confidence to perform a behaviour.

### Study Limitations

The main limitation of this study was related to the study design. We conducted a cross-sectional study to assess student’s intention to donate blood. Whereas the findings of this study reflect the students’ intention to donate blood at the time of the survey, there could have been changes in the students’ perceptions towards blood donation. The second limitation was related to the outcome used in this study. The study assessed students’ intention to donate blood rather than their actual blood donation practice.

## Conclusion

Most students had an intention to donate blood at some point in their life. The TPB was a weak predictor of the student’s intention to donate blood. Our study found that perceived behavioural control was main determinant of intentions to donated blood. Subjective norms and attitudes towards blood donation did not significantly influence the students’ intentions to donate. This could imply that for altruistic behaviours such as blood donation, attitudes towards donating blood do not matter but rather the persons’ perception of their ability to donate blood. Interventions should therefore be designed to make the internal and external environment of the students favourable for the students to donate blood.

## Data Availability

Data cannot be shared publicly because it consists of personal data. Pseudonymized will be available from the authors for researchers who clearly state the reason for which data is being requested.

## Acknowledgements

We would like to acknowledge the teachers and head teachers of Bunya secondary school, Jinja secondary school, Delta High School and St. James’ secondary school for giving use permission to conduct this study and for all the support rendered during the implementation of the study. We also acknowledge the blood donation champions in all the schools who worked with us to educate other students on the different aspects of blood donation. We also appreciate the students who took part in the study and completed both the baseline and end-line surveys.

## References

1. WHO. World Blood Donor Day: Be there for someone else. Give blood. Share life. Online2018 [5th November, 2023]. Available from: https://www.who.int/campaigns/world-blood-donor-day/2018.

2. Jenny HE, Saluja S, Sood R, Raykar N, Kataria R, Tongaonkar R, et al. Access to safe blood in low-income and middle-income countries: lessons from India. BMJ Global Health. 2017;2(2):bmjgh-2016-000167.

3. Dhingra N. World blood donor day: new blood for the world. World Health Organization. 2013.

4. WHO. Blood safety and availability. Fact sheet. Online2023 [6th November, 2023]. Available from: https://www.who.int/en/news-room/fact-sheets/detail/blood-safety-and-availability.

5. Bloch EM, Vermeulen M, Murphy E. Blood transfusion safety in Africa: a literature review of infectious disease and organizational challenges. Transfusion medicine reviews. 2012;26(2):164–80.

6. Zanin TZ, Hersey DP, Cone DC, Agrawal P. Tapping into a vital resource: Understanding the motivators and barriers to blood donation in Sub-Saharan Africa. African Journal of Emergency Medicine. 2016;6(2):70–9.

7. Okiror S,, January 16. . Doctors in Uganda Warn ‘crisis Level’ Blood Shortage Is Putting Lives at Risk. Online: The Guardian.; 2018 [18th November, 2023]. Available from: https://www.theguardian.com/global-development/2018/jan/16/doctors-uganda-warn-crisis-level-blood-shortage-risks-lives#:~:text=Uganda%20is%20grappling%20with%20a,to%20hospitals%2C%20is%20practically%20empty.

8. CDC. Global Health Initiative Executive Director Opens CDC-Supported Uganda Blood Transfusion Service Headquarters Online: Centers for Disease Control and Prevention; 2010 [6th November, 2023].

9. Murtagh CM, Katulamu C. Motivations and deterrents toward blood donation in Kampala, Uganda. Social Science & Medicine. 2021;272:113681.

10. Kassie A, Azale T, Nigusie A. Intention to donate blood and its predictors among adults of Gondar city: Using theory of planned behavior. PloS one. 2020;15(3):e0228929.

11. Uganda I. Knowledge, attitudes, and practices about regular, voluntary nonremunerated blood donation in Peri-urban and rural communities in Mbarara District, South Western Uganda, and its Impact on Maternal Health. J Obstet Gynaecol Can. 2015;37(10):903–4.

12. Godin G, Conner M, Sheeran P, Bélanger-Gravel A, Germain M. Determinants of repeated blood donation among new and experienced blood donors. Transfusion. 2007;47(9):1607–15.

13. Schlumpf KS, Glynn SA, Schreiber GB, Wright DJ, Randolph Steele W, Tu Y, et al. Factors influencing donor return. Transfusion. 2008;48(2):264–72.

14. Ajzen I. The theory of planned behavior. Organizational behavior and human decision processes. 1991;50(2):179–211.

15. Ajzen I. The theory of planned behaviour: Reactions and reflections. Taylor & Francis; 2011. p. 1113–27.

16. Ajzen I. From intentions to actions: A theory of planned behavior. Action control: From cognition to behavior: Springer; 1985. p. 11–39.

17. Wise D, Goggin KJ, Gerkovich MM, Metcalf KA, Kennedy SL. Predicting intentions to use condoms using gender, sexual experience, and the theory of planned behavior. American Journal of Health Education. 2006;37(4):210–8.

18. Huang M, Chen I, Chung S. The Theory of Planned Behavior for the Improvement of the Delayed Blood Donation Cycle, Optimization of the Planning Behavior, and Donor Intention. BioMed Research International. 2022;2022.

19. Abd Hamid NZ, Basiruddin R, Hassan N. Factors influencing the intention to donate blood: The application of the theory of planned behavior. International Journal of Social Science and Humanity. 2013;3(4):344.

20. Parash MH, Suki N, Shimmi S, Hossain A, Murthy K. Examining students’ intention to perform voluntary blood donation using a theory of planned behaviour: a structural equation modelling approach. Transfusion Clinique et Biologique. 2020;27(2):70–7.

21. Kiwanuka SN, Akulume M, Nankya FR, Kisakye AN. Evaluating the effect of targeted knowledge sharing on blood donation awareness and practices among secondary school students: A quasi-experimental study in Eastern Uganda. 2024.

22. Hossain MS, Siam MHB, Hasan MN, Jahan R, Siddiqee MH. Knowledge, attitude, and practice towards blood donation among residential students and teachers of religious institutions in Bangladesh–A cross-sectional study. Heliyon. 2022;8(10).

23. . Conceição VMd, Araújo JS, Oliveira RAAd, Santana MEd, Zago MMF. Perceptions of donors and recipients regarding blood donation. Revista brasileira de hematologia e hemoterapia. 2016;38:220–4.

24. Aschale A, Fufa D, Kekeba T, Birhanu Z. Intention to voluntary blood donation among private higher education students, Jimma town, Oromia, Ethiopia: Application of the theory of planned behaviour. PLoS One. 2021;16(3):e0247040.

25. Faqah A, Moiz B, Shahid F, Ibrahim M, Raheem A. Assessment of blood donation intention among medical students in Pakistan–An application of theory of planned behavior. Transfusion and Apheresis Science. 2015;53(3):353–9.

26. Gbadamosi FI, Popoola Y, Olaniyan F, Adesola RO, Unim B. Rural-urban variation in willingness to donate blood in Ibadan Region, Nigeria. Annali dell’Istituto Superiore di Sanita. 2023;59(2):114–21.

